# The Evolving Epidemiology of Infective Endocarditis: An Observational Study from Western India

**DOI:** 10.1101/2024.08.08.24311684

**Authors:** Surabhi Madan, Aditya Vyas, Dharshni Ramar, Manish Rana

**Affiliations:** Department of Infectious Diseases, Marengo CIMS Hospital, Ahmedabad, India; Department of Clinical Research, Marengo CIMS Hospital, Ahmedabad, India; Department of Community Medicine, GMERS Medical College, Sola, Ahmedabad, India

## Abstract

**Background:** Over the last few decades, the epidemiology and the management of infective endocarditis (IE) is undergoing significant evolution and advancements respectively. Many studies from the developed world and some from India have brought this to light.

**Objective:** To study demography, risk factors, microbiological spectrum, clinical profile, management, and outcome of IE in a tertiary care setup in western India.

Materials and Methods: This is a retrospective observational study of 90 patients diagnosed with definite as well as possible IE and admitted to the intensive care units (ICU) and wards in our institute from April 2015 to May 2022. Demographics, signs and symptoms, laboratory results, echocardiography, radiological findings, treatment, and outcomes were studied.

**Results:** 73 (81.1%) were definite and 17 (18.9%) were possible cases of IE. 93.3% cases were community-acquired, 6.7% were hospital acquired. Mean age of the patients was 43.3 + or - 18.8 years. Male: female ratio was 4.3:1. 62.2% had an underlying cardiac illness, with congenital heart disease (CHD) being the most common (18.9%) followed by rheumatic heart disease (RHD) (16.5%). Fever, weight loss, dyspnea, and cough were the common presenting symptoms. 69 (76.7 %) patients had native valve IE, 13 (14.4%) had prosthetic valve IE, 6 (6.7%) had vegetations on the endothelium without involvement of the valves, 2 (2.2%) patients had vegetations on the prosthetic material used for repair of congenital heart defects. About 48% of patients had a history of prior receipt of antibiotics. Blood cultures were positive in 48 (53.3%) patients. Most common valve infected was the mitral valve (MV) and the most common organisms isolated were the viridans group of streptococci (VGS), with penicillin (Pn) MIC <0.5 mcg/ml of all but one isolate. Transesophageal echocardiography (TEE), done in 17 patients, improved the pick-up of abnormal echo findings from 83.3% to 91.1%. Ceftriaxone was the most common antibiotic used. Surgery was done in 22 (24.4%) patients. 67 (74.4%) patients were cured, 14 (15.6%) patients died and 9 (10%) patients were lost to follow-up.

**Conclusion:** The increasing age of patients, absence of any underlying cardiac ailment, the importance of TEE in patients with high suspicion of IE, and the trend towards early surgery were the important aspects highlighted with respect to the changes happening in the field of IE. Almost all the strains of VGS, the most common isolated organisms, retained their sensitivity to Pn and ceftriaxone, making the treatment possible with once-a-day dosing of ceftriaxone monotherapy, highlighting the utilization of outpatient antibiotic therapy (OPAT).

## Introduction

Infective endocarditis (IE) is a rare, life-threatening infection of the endothelium of the heart (1). IE remains to be unchanged in its complexity and severity with high mortality and morbidity rates despite the advancements in diagnostics and overall healthcare (2, 3). Even in the modern era of medicine, IE with its ever-evolving nature, has an approximate incidence of 1.7–6.2 cases/100 000 patient years worldwide (3).

Multiple developments have occurred in the care of patients with IE including improvements in diagnostic capabilities, identification of patients eligible for OPAT and advances in the management of complications and risk-stratification of patients suffering from complications of IE, in particular stroke (4).

Compared to the western world, the epidemiology, clinical profile, and microbiologic spectrum of IE is different in India (5). Though the causes of IE in the western part of the world have now shifted to be largely due to intravenous (IV) drug abuse, presence of prosthetic valves, hemodialysis and presence of intra-cardiac devices (6); in India, many studies have found traditional risk factors such as RHD and CHD as the leading causes of IE (2, 5, 7, 8, 9).

The most commonly isolated etiological micro-organisms are VGS, staphylococci, enterococci, and fastidious gram-negative coccobacilli (2). *Staphylococcus aureus* has emerged as the most common cause of IE in most of the industrialized world (10), and the same has been reported in some Indian studies (7, 11).

This study was carried out to understand the current trends in the epidemiology of IE in western India.

## Methodology

The study period was from April 2015 to May 2022. This is a retrospective observational study of 90 patients diagnosed with IE and admitted to the intensive care units (ICU) and wards of a single tertiary care center in western India, whose files could be retrieved from the medical records department of the hospital. Patients of IE who were treated on outpatient basis were not included in the study.

Patients of all ages who were given the diagnosis of IE (definite and possible) were enrolled. Patient information was collected from the medical record files and the hospital’s intranet in a pre-designed semi-structured tool, pertaining to the demographics, signs and symptoms, laboratory results, echocardiography, radiological findings, treatment, and outcome.

Data was entered in google form and exported into MS office Excel. Data was analyzed using frequency distribution tables, cross tables; and proportions were calculated for appropriate variables.

The study was approved by the CIMS (Care Institute of Medical Sciences) hospital ethics committee.

## Results

During the study period, 90 patients who were diagnosed with IE were enrolled in the study. According to the Modified Duke’s criteria (12), 73 patients (81.1%) were labelled as definite cases of IE and 17 patients (18.9%) were categorized as “possible” cases. 93.3% of the patients acquired IE from the community, while 6.7% acquired it from the hospital. The characteristics of these 90 patients with IE are described in table 1. The mean age of the patients was 43.3 ± 18.8 years. The highest number of patients belonged to the 31-45 years age group (26.7%), followed by patients who were >60 years (23.3 %), 45-60 years (22.2%), 16-30 years (21.1%) and 0-15 (6.7%). The ratio of males: females was 4.3:1. 56 out of 90 patients (62.2%) had an underlying cardiac illness. The most common was presence of CHD (18.9%) followed by RHD (16.5%).

**Table 1:**
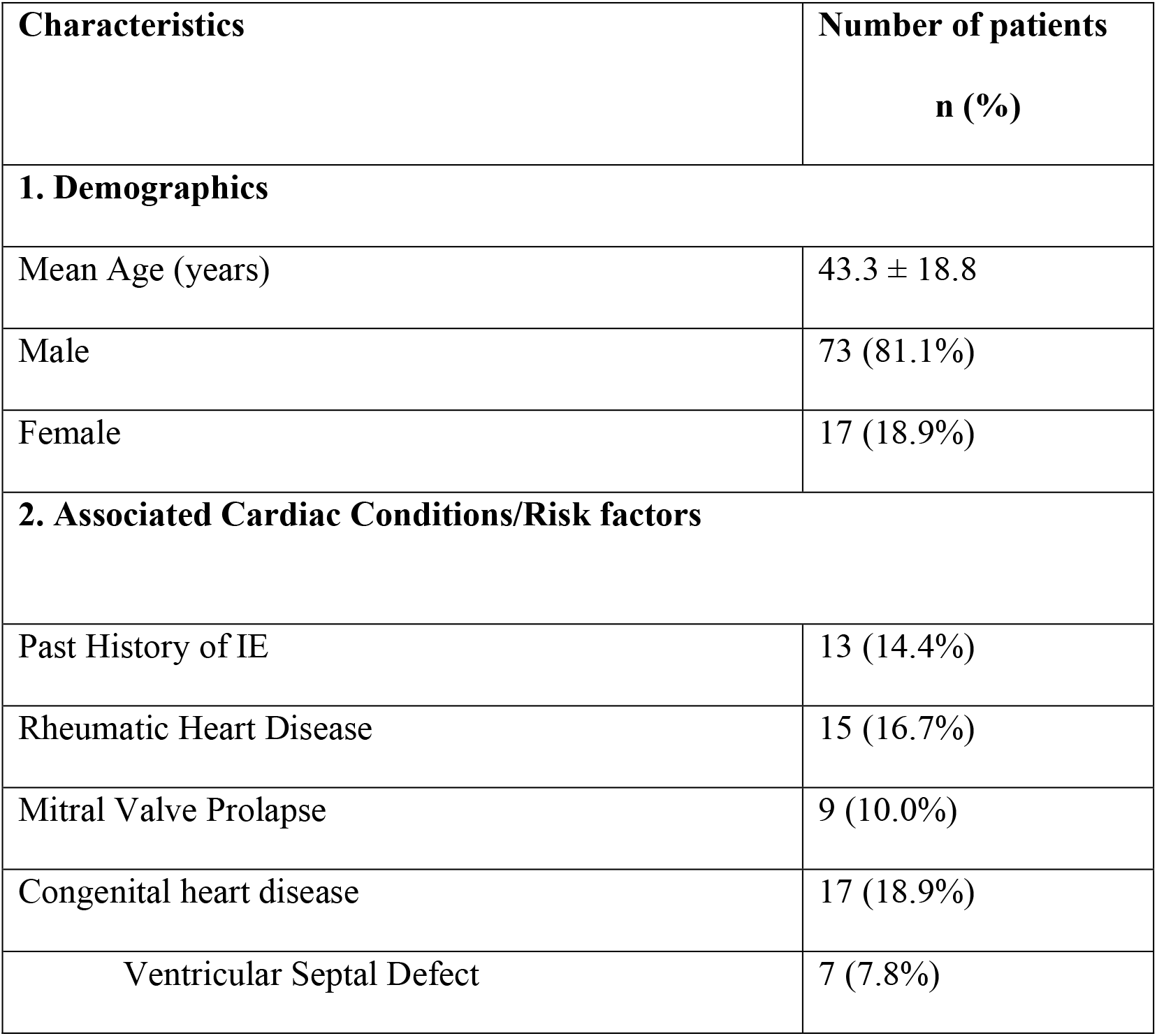
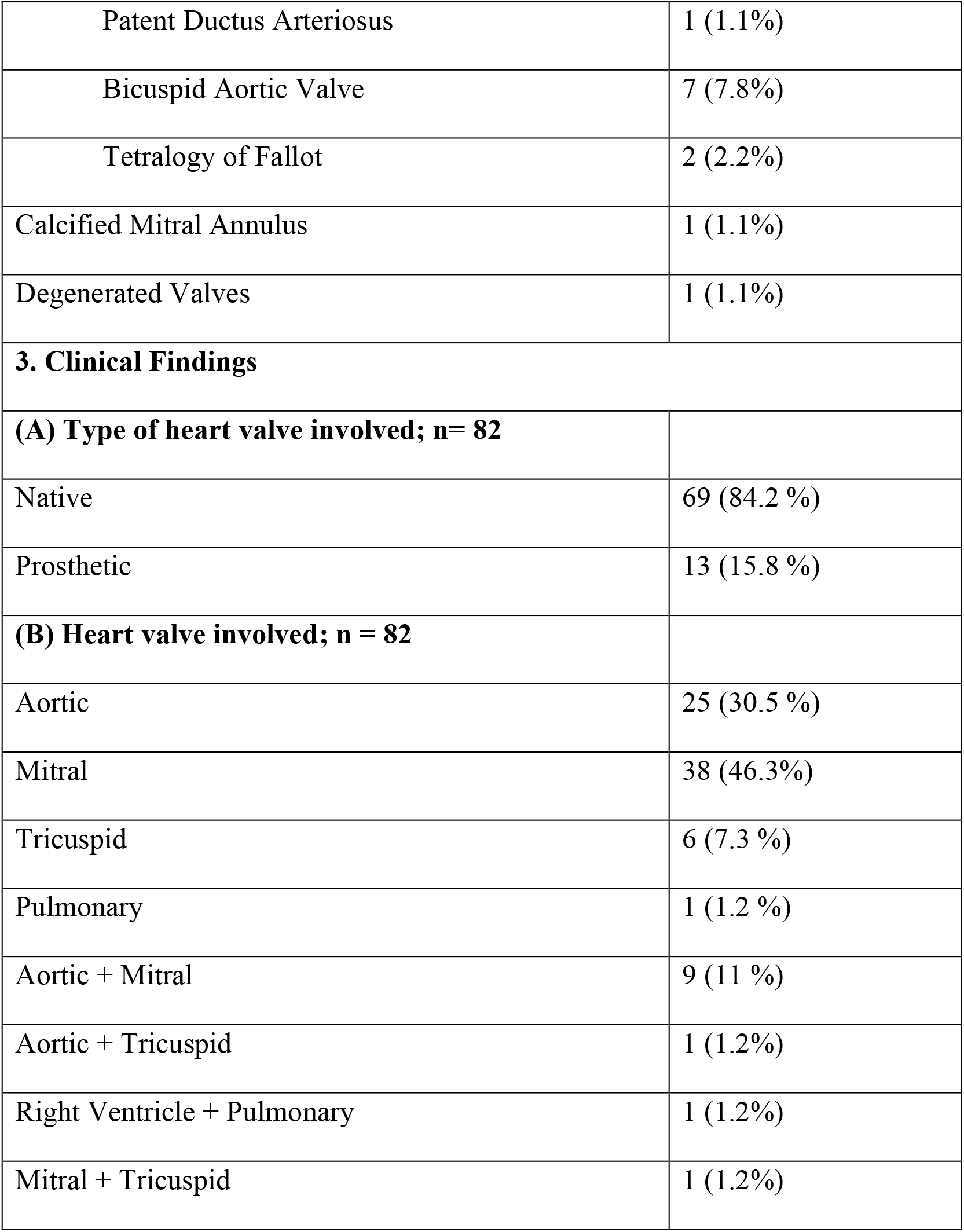
Characteristics of IE patients.

Involvement of a heart valve on echocardiography was found in 82 patients (91.1%). 69/90 (76.7 %) patients had native valve vegetations and 13/90 had involvement of prosthetic valve (14.4%). 6 (6.7%) patients had presence of vegetations on the endothelium without involvement of the valves. 2 (2.2%) patients had presence of vegetations on the prosthetic material / graft used for repair of congenital heart defects. Most commonly infected valve was the mitral valve (MV; 46.3%). In patients with RHD as well, MV was the most commonly affected valve.

Comorbidities present in these patients were hypertension (18.9%), diabetes mellitus (12.2%), coronary artery disease (8.9%), angioplasty (3.3%), coronary artery bypass grafting (5.6%), left ventricular dysfunction (8.9%), chronic kidney disease (5.6%), chronic liver disease (2.2%), tuberculosis (2.2%) and hypothyroidism (3.3%). HIV infection, rheumatoid arthritis, bronchial asthma was present in 1 patient each while one female was pregnant.

Clinical features are presented in table 2. 13.3% patients presented with various neurological symptoms like altered mental status, seizures, headache, ataxia, hemiparesis and decreased vision, slurred speech, sensory loss.

**Table 2:** Clinical features of patients with IE.

| Presenting symptoms | n (%) |
| --- | --- |
| Fever | 76 (84.4%) |
| Weight Loss | 38 (42.2%) |
| Dyspnea | 39 (43.3%) |
| Cough | 32 (35.6%) |
| Chest Pain | 15 (16.7%) |
| Abdominal pain | 14 (15.6%) |
| Nausea/vomiting | 13 (14.4%) |
| Neurological Symptoms | 12 (13.3%) |
| Palpitation | 6 (6.7%) |
| Diarrhea | 6 (6.7%) |
| Oliguria | 5 (5.6%) |

Echocardiography findings showed the presence of vegetation in 91.1% of all the patients. Valvular regurgitation was seen in 59 patients (65.6%), valvular stenosis in 14 patients (15.6%) and valvular/paravalvular leak in 5 patients (5.6%).

Transesophageal Echocardiography (TEE) was performed in 17 patients either to confirm the presence of vegetation or to rule out local complications. Performing TEE improved the percentage of abnormal echo findings with respect to presence of vegetation from 83.3% to 91.1%.

43/90 patients had history of receipt of antibiotics, most commonly, oral and injectable beta lactam antibiotics, cephalosporins, macrolides and fluoroquinolones; prior to the diagnosis of IE. Blood cultures were sent in all the patients. 3 sets (6 bottles) were sent in 49/90 patients. The details regarding the number of blood culture bottles collected and culture results are shown in table 3.

**Table 3:** Details of blood cultures in patients of IE.

| No. of Blood culture bottles sent | No. of patients | No. of patients with positive blood culture | No. of patients with negative blood culture |
| --- | --- | --- | --- |
| 2 | 3 | 3 | 0 |
| 3 | 5 | 3 | 2 |
| 4 | 21 | 11 | 10 |
| 5 | 3 | 2 | 1 |
| 6 | 49 | 23 | 26 |
| 7 | 1 | 0 | 1 |
| 8 | 2 | 2 | 0 |
| 9 | 6 | 4 | 2 |
|  | 90 | 48 | 42 |

Blood cultures were positive in 48 patients (53.3%). The most commonly isolated causative organisms (table 4) were VGS; the various species being *Streptococcus oralis, Streptococcus sanguinis, Streptococcus mitis, Streptococcus mutans, Streptococcus salivarius* and *Streptococcus gordonii*. Pn MICs were checked in 17 isolates of Streptococci; 8 (47.1%) were highly Pn sensitive (MIC ≤0.12 μg/ml), 8 isolates were relatively resistant to Pn (MIC >0.12 to <0.5 μg/ml) and 1 isolate had an MIC of 4 μg/ml.

**Table 4:** Organisms isolated in blood culture.

| **Organism Isolated (n = 48)** | **Native (n=38)** | **Prosthetic (n=10)** | **Total (n = 48)** |
| --- | --- | --- | --- |
| Viridans group of streptococci | 15 (31.2%) | 2 (4.2%) | 17 (35.4%) |
| *Streptococcus gallolyticus*<br>(*bovis*) | 6 (12.5%) | 0 (0) | 6 (12.5%) |
| *Enterococcus faecalis* | 6 (12.5%) | 1 (2.1%) | 7 (14.6%) |
| *Staphylococcus aureus* | 6 (12.5%) | 1 (2.1%) | 7 (14.6%) |
| Coagulase Negative<br>Staphylococci<br>(CONS) | 1 (2.1%) | 2 (4.2%) | 3 (6.3%) |
| *Pseudomonas aeruginosa* | 0 (0) | 2 (4.2%) | 2 (4.2%) |
| *Escherichia coli* | 1 (2.1%) | 1 (2.1%) | 2 (4.2%) |
| *Granulicatella adiacens* | 1 (2.1%) | 0 (0) | 1 (2.1%) |
| *Burkholderia cenocepacia* | 1 (2.1%) | 0 (0) | 1 (2.1%) |
| *Salmonella typhi* | 0 (0) | 1 (2.1%) | 1 (2.1%) |
| *Brucella melitensis* | 1 (2.1%) | 0 (0) | 1 (2.1%) |

Majority of the patients of VGS endocarditis and all patients with IE due to *Streptococcus gallolyticus* had native valve involvement. On the other hand, both the patients with IE due to *Pseudomonas aeruginosa* had PV IE. Amongst the patients with culture positive PV IE, the most common organisms were CONS (20%), VGS (20%) and *Pseudomonas aeruginosa* (20%).

The most common complications seen in the study were cardiac failure (18.1 %) followed by neurological complications (15.6%), which included cerebral infarcts, embolism and hemorrhage. Other complications were pulmonary embolism, peripheral arterial embolism, splenic infarct, acute kidney injury, sepsis.

The most common antibiotics used for treatment were ceftriaxone monotherapy (34.4%) followed by ampicillin and ampicillin-sulbactam; both in combination with gentamicin/ceftriaxone/vancomycin (30%). Less commonly used antibiotics used were cefazolin, ceftazidime, cefoperazone-sulbactam, meropenem, imipenem, cloxacillin, vancomycin, teicoplanin, daptomycin, linezolid, cotrimoxazole and ciprofloxacin.

Table 5 shows the treatment modalities and the final outcomes. 68 patients (75.6%) required only medical treatment while surgical intervention was required in 22 patients (24.4%). Data for the timing of surgery was available for 16 (out of 22) patients. 3 patients were operated within 2 weeks of diagnosis of IE while 8 patients got operated in 2 – 4 weeks period after the diagnosis. 5 patients were taken for surgery after 4 weeks of IE diagnosis.

**Table 5:** Treatment modalities and outcomes in patients with IE.

| Organism Isolated (n = 48) | Treatment Modality |  | Outcomes |  |  | Total |
| --- | --- | --- | --- | --- | --- | --- |
|  | Medical Treatment | Medical Treatment with surgery | Cured n (%) | Died n (%) | Lost to follow up n (%) |  |
| Viridans group of streptococci | 12 (70.6) | 5 (29.4) | 13 (76.5) | 4 (23.5) | 0 | 17 |
| *Streptococci gallolyticus (bovis)* | 5 (83.3) | 1 (16.7) | 5 (83.3) | 1 (16.7) | 0 | 6 |
| *Enterococcus faecalis* | 5 (71.4) | 2 (28.6) | 4 (57.1) | 3 (42.9) | 0 | 7 |
| *Staphylococcus aureus* | 6 (85.7) | 1 (14.3) | 6 (85.7) | 1 (14.3) | 0 | 7 |
| Coagulase Negative Staphylococci (CONS) | 1 (33.3) | 2 (66.7) | 3 (100) | 0 | 0 | 3 |
| *Pseudomonas aeruginosa* | 2 (100) | 0 | 2 (100) | 0 | 0 | 2 |
| *Escherichia coli* | 2 (100) | 0 | 1 (50) | 1 (50) | 0 | 2 |
| *Granulicatella adiacens* | 1 (100) | 0 | 1 (100) | 0 | 0 | 1 |
| *Burkholderia cenocepacia* | 1 (100) | 0 | 0 | 0 | 1 (100) | 1 |
| *Salmonella typhi* | 1 (100) | 0 | 1 (100) | 0 | 0 | 1 |
| *Brucella melitensis* | 0 | 1 (100) | 1 (100) | 0 | 0 | 1 |
| Culture negative | 32 (76.2) | 10 (23.8) | 30 (71.4) | 4 (9.5) | 8 (19.1) | 42 |
| Total | 68 (75.6) | 22 (24.4) | 67 (74.4) | 14 (15.6) | 9 (10) | 90 |

67 (74.4%) patients were cured while 14 patients died (15.6%) and 9 (10%) patients were lost to follow-up.

## Discussion

There is substantial evidence from the western world towards significant change in the epidemiology of IE (10). We studied and analyzed the epidemiological characteristics in patients presenting to a tertiary care hospital of western India.

Most of our patients were above 40 years of age, which is similar to a study from north India (7), where the mean age of patients was 49.3 ± 13.7 years. While this is similar to studies conducted in the western world, (12, 13) it is in contrast to the studies conducted earlier in India (14, 15, 16), in which the mean age was much lower. The possible reasons could be the decreasing proportion of rheumatic heart disease patients, and the aging Indian population with access to the recent advancements in diagnostic and therapeutic methods.

In our study, the most common risk factor was CHD followed by RHD, which again highlights a slow paradigm shift in the epidemiology of IE in India.

The clinical presentation and complications were similar to studies done in the past, with fever and congestive cardiac failure as the most common presenting symptom and complication respectively (7, 14). Our overall echocardiographic detection rate of vegetation was 91.1%. TEE improved the percentage of abnormal echo findings with respect to the presence of vegetation, from 83.3% to 91.1%. This is in accordance with recent reports on the sensitivity of TEE (17).

In the present study, the culture positivity was found to be 53.3%, which is in contrast to culture positivity reports of 69 to 97% in the west. (12) Prior antibiotic use before proper diagnostic workup is the most common responsible factor (7).

VGS were the most common organisms in our study, which is similar to previous studies in India (7, 16) though there are studies with *Staphylococcus aureus* as the most common etiological agent for IE (8). The majority of patients with VGS endocarditis and all patients who grew *Streptococcus gallolyticus* had native valve involvement. In contrast, both the patients with IE due to *Pseudomonas aeruginosa* had PVE. Among patients with culture-positive PVE, the most common organisms were CONS and VGS, like other studies (18). Almost all the isolates of VGS were found to have Pn MIC <0.5, making treatment with ceftriaxone monotherapy as the most common regimen for treatment.

The overall in-hospital mortality rate in the present study was 15.6%. Similar studies have reported an overall decrease in mortality due to IE in India of 42% in 1970 (19), 25% in 1992 (14), 13% in 2001 (20), and 6.5% in 2013 (7). The relatively high mortality rate in our study could be attributed to the fact that ours is a tertiary care center that receives relatively complicated cases.

## Limitations

Our major limitation was that this was an observational, retrospective, single-center study. The diagnostic work up and treatment was not protocol driven and was done as per consultants’ discretion. Molecular and serological investigations were not utilized fully due to limitations of access to the tests and cost. We could not assess the outcomes with respect to the timing of the surgery. 10% of the patients were lost to follow-up.

## Conclusion

This study reinforced the evolutionary phenomenon happening in the epidemiology of IE. Increasing age, absence of any underlying cardiac ailment, importance of TEE in patients with high suspicion of IE, and trend towards early surgery were noticed in this study. Prior administration of antibiotics by the primary care doctors, before establishing the diagnosis of IE and before sending blood cultures, is a major factor towards poor sensitivity of blood cultures, and hence possible ‘overuse’ of antibiotics. Almost all the strains of VGS, the most common organisms isolated, retained their sensitivity to penicillin and ceftriaxone, making the treatment possible with once-a-day dosing of ceftriaxone monotherapy, highlighting the utilization of outpatient antibiotic therapy (OPAT).

## Data Availability

All data produced in the present work are contained in the manuscript

## Notes

### Author Contributions

Surabhi Madan conceived and designed the study. Surabhi Madan and Dharshni Ramar wrote the manuscript. Aditya Vyas collected the and entered the data in MS excel. Manish Rana performed the analysis and reviewed the manuscript. All authors read and approved the final manuscript.

## ACKNOWLEDGMENT

The authors thank Dr Riddhi Parikh, Dr Disha Patel, Department of Clinical Research, Marengo CIMS Hospital, Ahmedabad, India, for their assistance in data aggregation.

